# Usability and Physician Acceptance of AI in Chest Radiography: An Evaluation of the MeDeep Platform Using SUS and ATRAI-14

**DOI:** 10.64898/2026.10.23.26365338

**Authors:** Parand Shariat Rad, Mohammad Mehdi Hatami, Mohammad Amin Amani, Shayan Salehi, Mohammad Hossein Sharif, Mohammad Javad Gharegozlou, Mohammad Gharib Salehi

## Abstract

**Objectives:** Artificial intelligence (AI) architectures demonstrate high proficiency in chest radiography interpretation, yet successful clinical integration is often hampered by usability challenges and physician skepticism. This study evaluates the ergonomic usability and professional acceptance of the MeDeep AI platform among medical practitioners to identify key drivers of clinical trust.

**Methods:** A multicenter, cross-sectional study was conducted across hospitals in Kermanshah, evaluating 88 medical professionals, including attending radiologists, radiology residents, internal medicine physicians, and medical externs. The System Usability Scale (SUS) was utilized to measure the platform’s ergonomic functionality (n = 82), while the validated ATRAI-14 questionnaire quantified broader attitudes and trust regarding AI integration (n = 88).

**Results:** The MeDeep platform achieved a high mean SUS score of 84.93, indicating excellent usability uniformly experienced across all professional subgroups (p = 0.387). However, overarching AI acceptance and algorithmic trust varied significantly by clinical role. Radiology residents exhibited significantly higher overarching AI acceptance (p = 0.016) and algorithmic trust (p = 0.008) compared to attending radiologists. Multivariate regression demonstrated that user familiarity (Beta = 0.500, p < 0.001) and perceived system usability (Beta = 0.236, p = 0.012)—rather than years of clinical experience—were the primary independent positive predictors of AI acceptance. Furthermore, higher perceived usability directly predicted greater algorithmic trust (Beta = 0.282, p = 0.009).

**Conclusion:** While technical usability is a necessary baseline for diagnostic platforms, professional trust dictates ultimate clinical adoption. Overcoming the generational divide in AI acceptance requires developers to prioritize intuitive interface design and position AI as a workflow-augmenting tool that addresses administrative burdens.

**Key Points:**

- The MeDeep AI platform demonstrated excellent ergonomic usability uniformly across all professional clinical ranks.
- Significant generational divides exist, with residents exhibiting higher algorithmic trust than attending radiologists.
- User familiarity and perceived usability predict overarching AI acceptance better than years of clinical experience.

## 1. Introduction

Chest radiography remains the most frequently performed imaging examination globally, serving as a cost-effective and essential frontline diagnostic tool for a vast spectrum of cardiopulmonary diseases [1, 2]. Despite its ubiquity, the interpretation of chest X-rays is highly complex and inherently vulnerable to human error. Unprecedented clinical demand has precipitated a surge in imaging volumes, culminating in severe professional burnout, fatigue, and an elevated risk of diagnostic latency among radiologists. These systemic pressures contribute to substantial inter-observer variation and high miss rates for critical, early-stage pathologies, acting as primary factors in the delayed diagnosis of conditions such as lung cancer [3-6].

To address these vulnerabilities and build greater resilience within healthcare systems, artificial intelligence (AI) has emerged as a transformative paradigm. Recent deep neural networks and advanced multimodal platforms have achieved radiologist-level proficiency in detecting complex thoracic pathologies, including pneumonia, pneumothorax, and pleural effusion [7-9]. By acting as an automated analytical aid, previously developed and validated AI architectures—such as the MeDeep platform utilized in this study—are specifically designed to reduce errors resulting from commonly missed radiographic findings. Emerging evidence from diverse clinical environments, ranging from emergency departments to field hospitals, indicates that AI-assisted interpretation can significantly augment physician decision-making and diagnostic precision [10-12].

However, the promising computational performance of these algorithms does not automatically translate to successful clinical integration. The deployment of medical AI is frequently hampered by poor system usability, a lack of trust in diagnostic explanations, and complex psychological barriers among end-users. Recent cross-sectional assessments reveal a deeply heterogeneous landscape of AI adoption among healthcare workers. While many physicians’ express optimism regarding AI’s potential to improve patient outcomes and alleviate reporting burdens, significant apprehensions persist regarding technological reliability, shifts in professional workload, and the ethical implications of autonomous decision-making. Bridging the gap between algorithmic accuracy and routine clinical utility requires a rigorous evaluation of human-computer interaction, organizational intelligence, and the broader service quality gaps that directly impact physician acceptance [13-15].

Consequently, this study aims to comprehensively conduct an independent evaluation of the established MeDeep chest radiography platform’s usability and physician acceptance. To achieve a robust, multidimensional assessment, this research employs the System Usability Scale (SUS) [16, 17] as a reliable, industry-standard metric for evaluating the perceived ease of use, efficiency, and learnability of digital health platforms. Concurrently, the validated ATRAI-14 questionnaire [18] is utilized to precisely quantify radiologists’ attitudes toward AI implementation across four critical domains: Familiarity, Trust, Implementation Perspectives, and Hopes and Fears. By mapping both the ergonomic usability and the complex attitudinal landscape of the physicians interacting with the MeDeep platform, this evaluation provides actionable insights to facilitate the seamless and effective adoption of AI in modern diagnostic imaging.

## 2. Methods

### Study Design and Setting

A multicenter, cross-sectional observational study was conducted across public and private hospitals in Kermanshah over a five-month period in 2025 to evaluate the clinical usability and professional acceptance of the MeDeep artificial intelligence platform. The study protocol was specifically designed to capture real-world human-computer interactions and the nuanced attitudes of medical practitioners utilizing AI for chest radiography interpretation independently of the platform’s original developers.

### Participants

The study cohort comprised medical professionals directly involved in diagnostic imaging and patient care. A total of 88 valid responses were recorded for the overarching attitudinal assessment, of which 82 participants fully completed the platform-specific usability evaluation. Six participants were excluded from the platform-specific usability evaluation due to incomplete responses but were retained for the broader attitudinal assessment. The demographic profile of the primary cohort yielded a mean age of 37.91 years, consisting of 63.41% male and 36.59% female practitioners. To ensure a diverse representation of clinical experience, the professional distribution was stratified into four distinct subgroups: attending radiologists (45.12%), radiology residents (36.59%), internal medicine physicians (13.41%), and medical externs (4.88%). Regarding prior technological exposure, 63.41% of participants reported having previous opportunities to utilize AI tools for interpreting imaging studies within their clinical workflow, whereas 36.59% were entirely naive to such diagnostic platforms.

### Instruments

Data were prospectively collected utilizing two distinct, validated psychometric instruments designed to measure micro-level usability and macro-level professional attitudes, respectively:

- **System Usability Scale (SUS):** The SUS, a standardized 10-item questionnaire, was employed to evaluate the ergonomic functionality, learnability, and perceived ease of use of the MeDeep interface. Within this cohort, the instrument demonstrated robust internal consistency, yielding a Cronbach’s alpha of 0.793.
- **ATRAI-14 Questionnaire:** The 14-item ATRAI-14 instrument was deployed to quantify practitioners’ broader perceptions regarding AI integration into the clinical environment. The questionnaire maps cognitive and professional attitudes across critical sub-domains, explicitly isolating factors such as Trust, Implementation Perspectives, and Hopes. This tool also exhibited acceptable reliability within the study sample, achieving a Cronbach’s alpha of 0.784.

### Statistical Analysis

All data preparation and statistical computing were executed using IBM SPSS Statistics software. Descriptive statistics were generated to summarize demographic variables, imaging modality expectations, and baseline professional characteristics. Survey reliability was confirmed utilizing Cronbach’s alpha for both psychometric instruments.

To examine mean differences among the four professional roles (radiologists, residents, internal medicine physicians, and externs), a one-way analysis of variance (ANOVA) was conducted, followed by Tukey’s Honestly Significant Difference (HSD) post-hoc testing to pinpoint specific group variances in trust and system usability. Bivariate relationships—particularly evaluating how prior clinical AI experience and overall usability scores correlated with general attitudes—were analyzed using Pearson and Spearman rank correlation coefficients. Finally, multiple linear regression modeling was employed to ascertain the predictive value of specific variables (namely, perceived system usability, user familiarity, and prior AI experience) on overarching physician trust and AI acceptance. The threshold for statistical significance was established *a priori* at the 0.05 level (2-tailed).

## 3. Results

### Participant Characteristics

A total of 88 medical professionals participated in the attitudinal assessment, with complete demographic and system usability data available for a primary sub-cohort of 82 individuals. Within this valid cohort (n = 82), the mean age was 37.91 years. The sample comprised 52 male (63.41%) and 30 female (36.59%) participants. Professional distribution included 37 attending radiologists (45.12%), 30 radiology residents (36.59%), 11 internal medicine physicians (13.41%), and 4 medical externs (4.88%). Furthermore, 52 participants (63.41%) reported prior experience utilizing artificial intelligence (AI) tools in their clinical workflows, whereas 30 individuals (36.59%) were entirely naïve to such technologies.

**Table 1:**
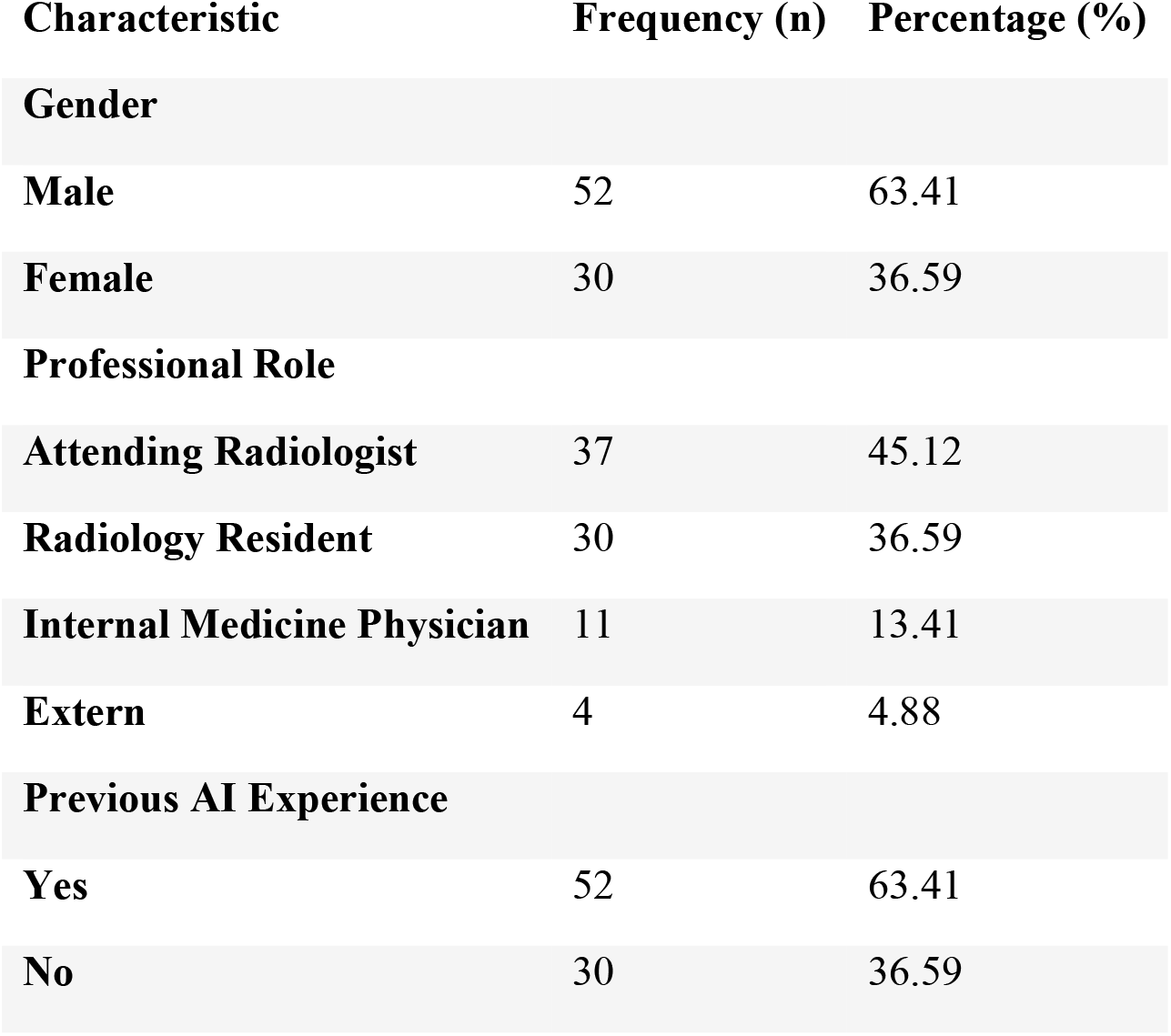
Baseline Demographic and Professional Characteristics (n = 82)

### Perceived Utility and Impact of AI

Participants anticipated that AI integration would most significantly impact radiography (n = 67), followed by computed tomography (n = 57), mammography (n = 56), and magnetic resonance imaging (n = 51). Conversely, 10 participants believed AI would have no meaningful impact on diagnostic imaging. Regarding clinical utility, the most anticipated functions of AI included assistance with structured reporting (n = 63), extraction of quantitative data such as size and volume (n = 58), and image post-processing for noise reduction (n = 51). Incidental finding detection (n = 49) and cancer staging (n = 48) were also identified as highly valuable clinical functions.

**Table 2:**
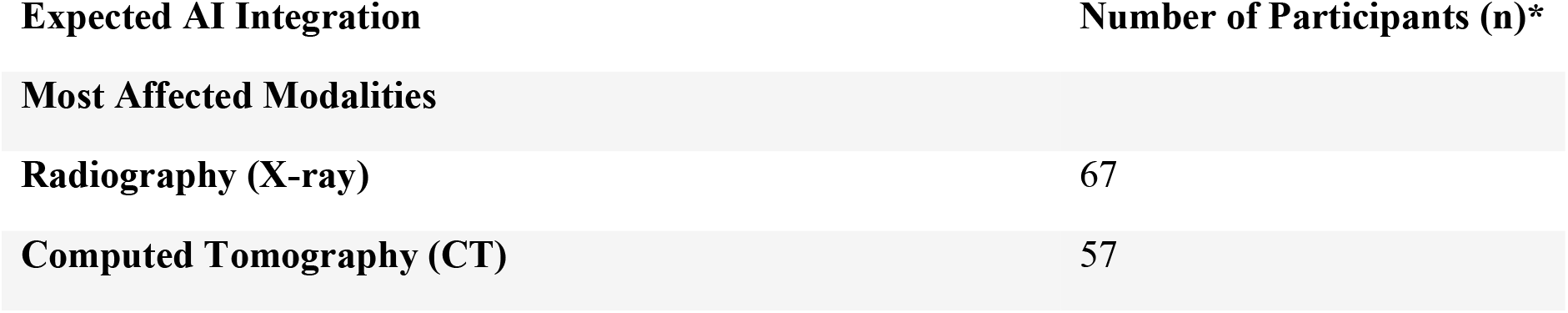
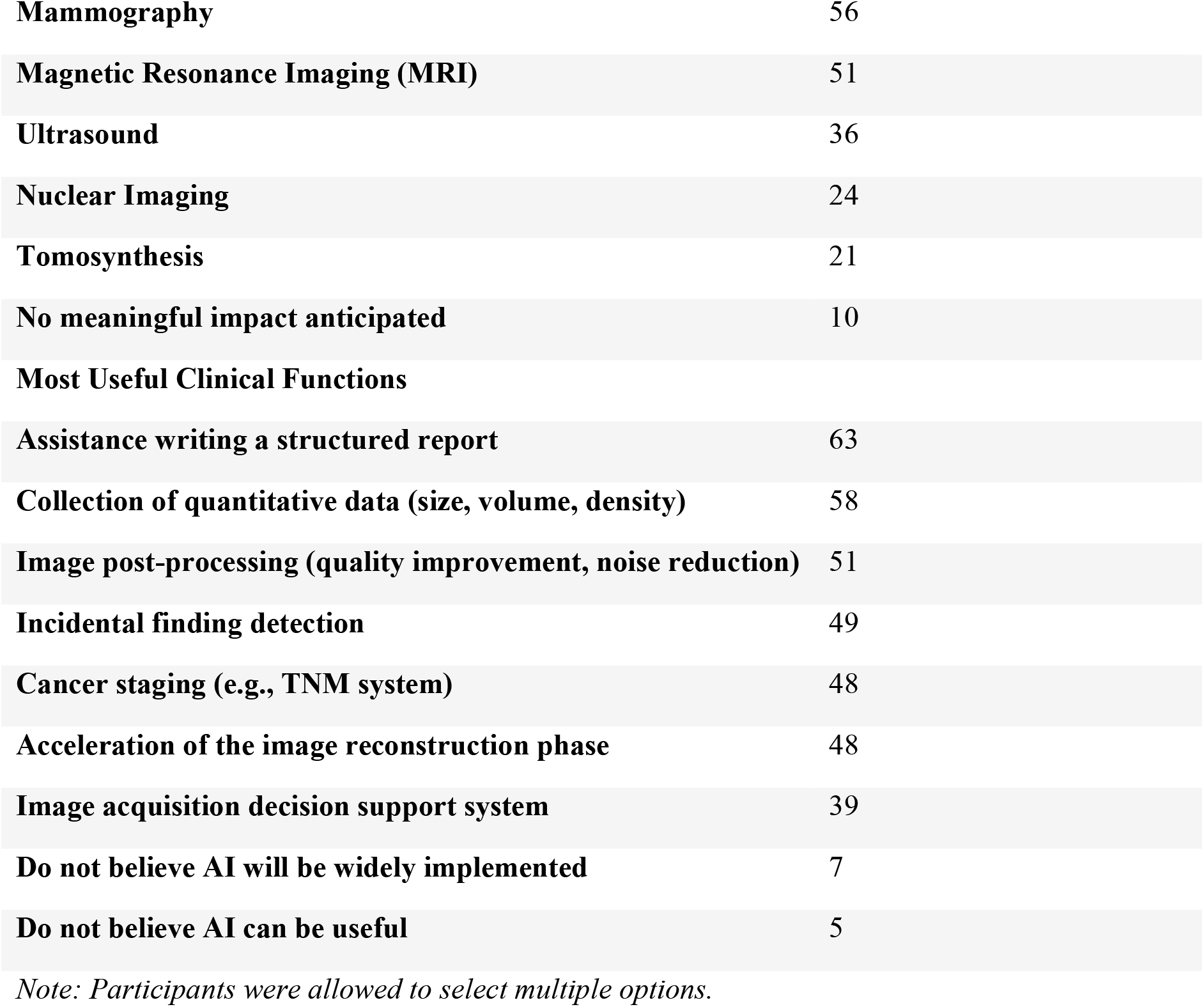
Perceived Impact and Clinical Utility of Artificial Intelligence.

### System Usability Scale (SUS) Evaluation

The MeDeep platform demonstrated excellent overall usability, achieving a mean SUS score of 84.93 (SD = 9.45; 95% CI: 82.85–87.00) among the 82 evaluating participants. The instrument maintained robust internal consistency with a Cronbach’s alpha of 0.793. A one-way analysis of variance (ANOVA) indicated no statistically significant differences in perceived usability across the four professional subgroups (F = 1.024, p = 0.387), confirming that the interface was uniformly accessible to all clinical ranks.

**Table 3:**
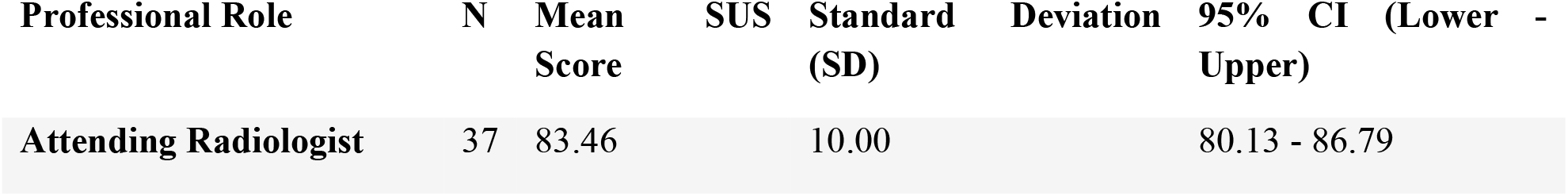
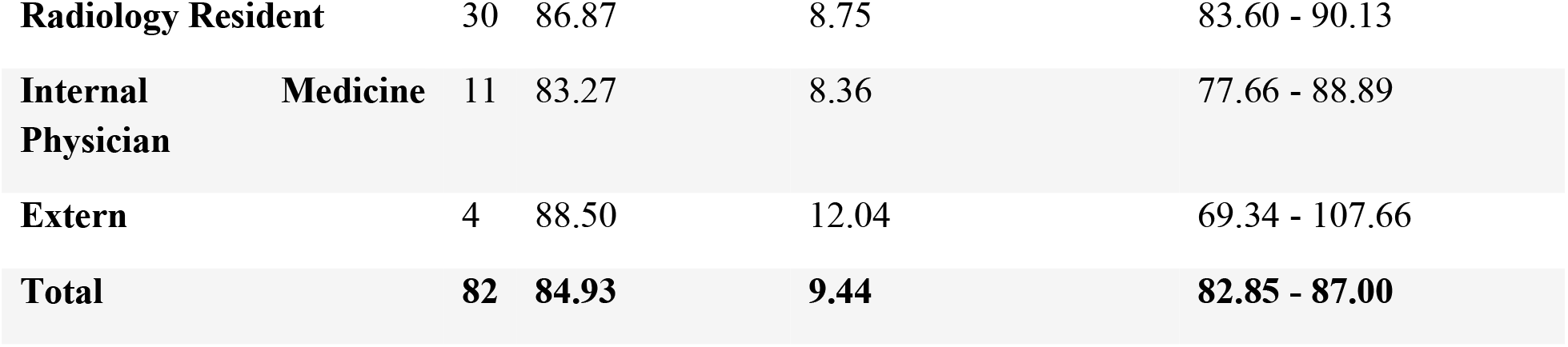
System Usability Scale (SUS) Scores by Professional Role.

### ATRAI-14 Assessment and Professional Acceptance

Broader attitudes toward AI implementation were assessed using the ATRAI-14 questionnaire across all 88 participants, demonstrating acceptable internal reliability (Cronbach’s alpha = 0.784). The mean total ATRAI-14 score was 21.35 (SD = 5.09; 95% CI: 20.27–22.43). ANOVA revealed statistically significant variance in overall AI acceptance among different professional roles (F = 2.681, p = 0.037). Tukey’s HSD post-hoc testing demonstrated that radiology residents (mean = 23.31) exhibited significantly more positive overarching attitudes toward AI compared to attending radiologists (mean = 19.54; p = 0.016).

This generational divergence was exceptionally pronounced within the “Trust” sub-domain (overall mean = 7.97, SD = 3.03). ANOVA confirmed significant group differences in trust (F = 3.065, p = 0.021), and post-hoc testing indicated that residents scored significantly higher in algorithmic trust (mean = 9.19) than their attending counterparts (mean = 6.82; p = 0.008).

**Table 4:**
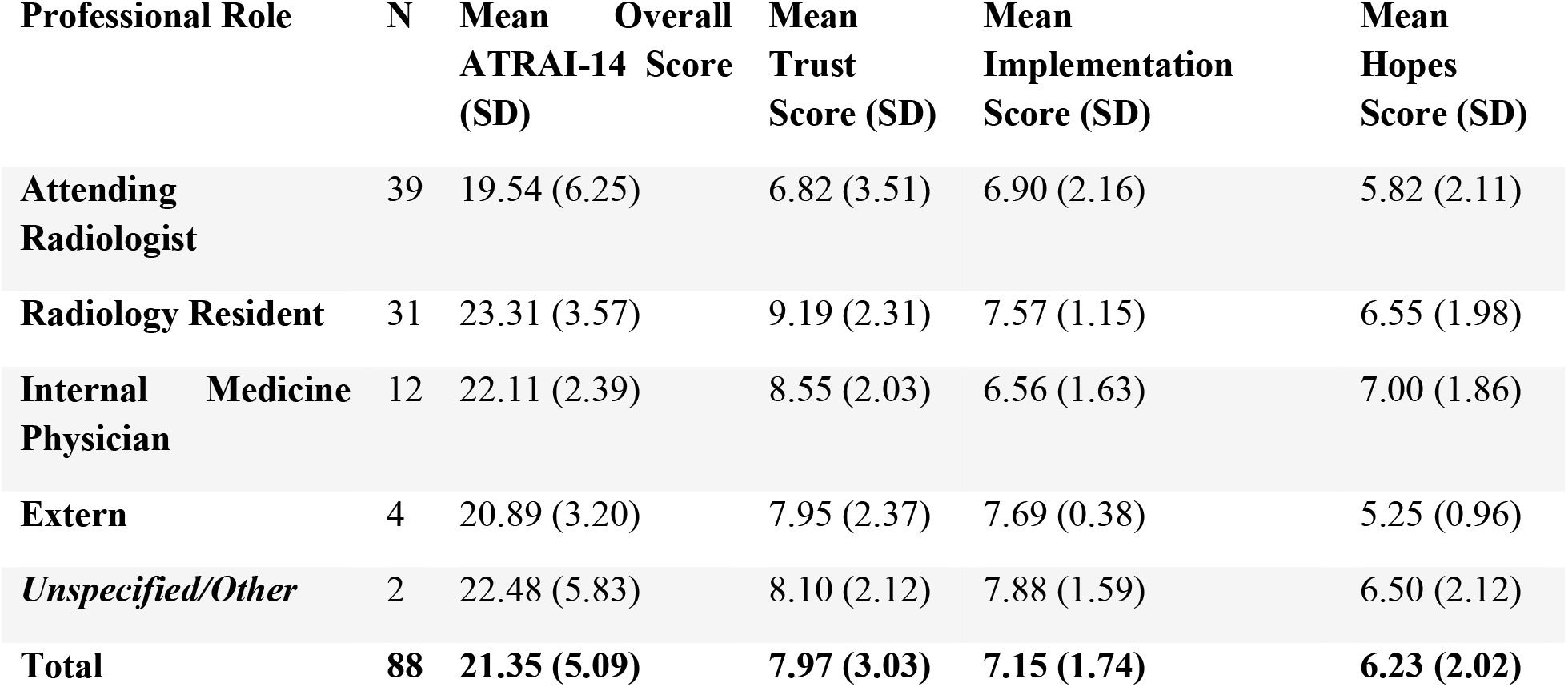
Overall ATRAI-14 and Trust Sub-domain Scores by Professional Role.

### Correlational and Regression Analysis

Bivariate correlation analyses demonstrated an inverse relationship between clinical experience and technological acceptance. Increased years of professional experience correlated significantly with lower overall ATRAI-14 scores (Pearson’s r = -0.262, p = 0.014) and slightly lower SUS scores (Spearman’s rho = -0.221, p = 0.046).

To ascertain the primary drivers of AI acceptance, a multiple linear regression model was constructed to predict the total ATRAI-14 score based on user familiarity, total SUS score, and years of clinical experience. The overall model was statistically significant (R^2^ = 0.350, Adjusted R^2^ = 0.326, F = 14.378, p < 0.001). Both familiarity with AI (Beta = 0.500, p < 0.001) and perceived system usability (Beta = 0.236, p = 0.012) emerged as significant positive independent predictors of overarching AI acceptance. In contrast, clinical experience did not retain independent predictive value in this multivariate model (p = 0.567). A secondary regression model confirmed that a higher perceived system usability directly and significantly predicted higher levels of trust in the AI platform (R^2^ = 0.080, Beta = 0.282, p = 0.009) (Table 5).

**Table 5:**
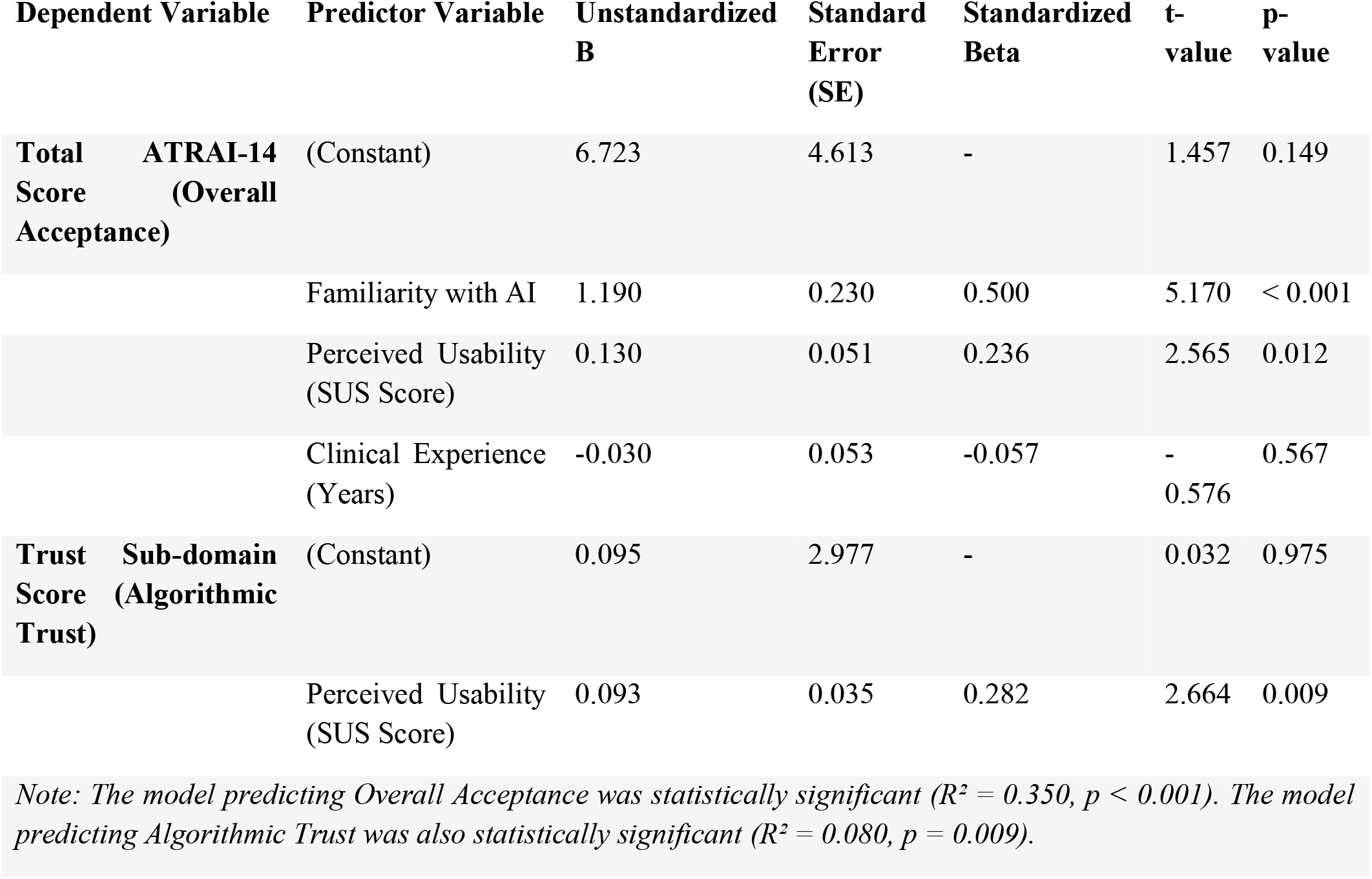
Linear Regression Models Predicting Overall AI Acceptance and Algorithmic Trust.

## 4. Discussion

### Principal Findings and Clinical Implications

The successful integration of artificial intelligence into diagnostic radiology requires bridging the formidable gap between algorithmic proficiency and human acceptance. This independent evaluation of the MeDeep platform yields a critical clinical insight: while technical usability is a necessary baseline, professional trust remains the ultimate arbiter of AI adoption. The MeDeep platform demonstrated exceptional ergonomic performance, achieving a mean System Usability Scale (SUS) score of 84.93. This score places the platform well above the established industry average of 68, indicating superior learnability and user satisfaction. Notably, this high usability was uniformly experienced across all professional roles, from medical externs to attending radiologists, with no statistically significant differences observed (p = 0.387). However, our findings robustly demonstrate that a diagnostic platform’s ease of use does not automatically guarantee uniform clinical trust.

### The Generational Divide in AI Trust and Acceptance

A central discovery of this study is the significant divergence in AI acceptance between professional tiers. Although attending radiologists found the MeDeep system just as easy to navigate as their junior counterparts, they exhibited significantly lower overarching acceptance (p = 0.016) and algorithmic trust (p = 0.008). This generational divide underscores a profound psychological barrier within the reading room. Radiology residents—who are largely digital natives—may inherently view AI as a collaborative, integrated tool designed to optimize workflow. In contrast, highly experienced attending physicians rely on deeply ingrained clinical heuristics, fostering a natural and protective skepticism toward autonomous, machine-driven decision-making.

This inverse relationship is further corroborated by our correlational analysis, which demonstrated that increased years of professional experience significantly correlated with lower overall ATRAI-14 scores (r = -0.262, p = 0.014). These findings align with recent literature suggesting that senior clinicians often prioritize explainability and diagnostic rationale over mere computational output. For AI to be successfully adopted by senior staff, developers must prioritize “explainable AI” (XAI) models that transparently delineate how an algorithm arrived at its conclusion, rather than presenting a “black box” diagnosis [19].

### Usability and Familiarity as Catalysts for Adoption

Crucially, our multivariate regression analysis offers a strategic roadmap for overcoming this professional skepticism. The data explicitly demonstrate that prolonged clinical experience alone does not dictate AI rejection; in the multivariate model, experience lost its independent predictive value (p = 0.567). Rather, user familiarity (Beta = 0.500, p < 0.001) and perceived system usability (Beta = 0.236, p = 0.012) emerged as the primary positive predictors of overarching AI acceptance.

Furthermore, our secondary regression model confirmed that higher ergonomic usability directly and significantly predicted higher levels of trust in the AI platform (R^2^ = 0.080, Beta = 0.282, p = 0.009). This outcome strongly supports the principles of the Technology Acceptance Model (TAM), suggesting that transparent, highly intuitive interfaces actively cultivate physician confidence over time [20].

Additionally, participants heavily favored utilizing AI for tasks that alleviate administrative burden, such as structured reporting (n = 63), quantitative data extraction (n = 58), and image post-processing (n = 51). Physicians are far more willing to adopt AI when it acts as a quantitative shield against burnout, augmenting human expertise rather than attempting to replace primary diagnostic functions [21, 22].

### Limitations and Future Directions

This study acknowledges certain methodological limitations. The cross-sectional design and specific regional focus within Kermanshah medical facilities may limit broader geographic generalizability. There was a slight attrition rate between the attitudinal assessment (n = 88) and the usability evaluation (n = 82), which may introduce minor selection bias, though the core demographic distributions remained stable. Additionally, self-reported psychometric assessments can be subject to response biases. Future longitudinal research should investigate whether prolonged, mandatory exposure to highly usable platforms like MeDeep actively normalizes trust among senior clinicians and directly correlates with a measurable reduction in actual diagnostic error rates.

## 5. Conclusion

The successful clinical integration of artificial intelligence in chest radiography extends beyond algorithmic accuracy, depending heavily on system usability and physician trust. This evaluation of the MeDeep platform demonstrates that while modern AI tools can achieve exceptional ergonomic usability across all levels of medical training, profound generational divides in algorithmic trust remain a significant barrier. Highly experienced attending physicians exhibit considerably more skepticism toward autonomous diagnostic aids compared to their resident counterparts. Crucially, this study reveals that user familiarity and intuitive interface design are the primary catalysts for overarching AI acceptance, effectively mitigating the hesitations associated with prolonged clinical experience. To facilitate widespread adoption and reduce diagnostic burnout, healthcare institutions and developers must position AI not as an autonomous replacement, but as a transparent, highly usable collaborative tool tailored to augment physician workflow and alleviate administrative burdens.

## Data Availability

The datasets generated and analyzed during the current study are not publicly available due to privacy restrictions but are available from the corresponding author on reasonable request.

## Abbreviations

AI: Artificial Intelligence
SUS: System Usability Scale
CT: Computed Tomography
MRI: Magnetic Resonance Imaging

## Declarations

### Ethics Approval and Consent to Participate

This study was performed in line with the principles of the Declaration of Helsinki. Approval was granted by the Ethics Committee of

Kermanshah University of Medical Sciences (No: IR.KUMS.MED.REC.1405.053). Informed consent was obtained from all individual participants included in the study.

### Consent for Publication

Not applicable.

## Competing Interests

The authors declare that they have no competing interests.

## Funding

The authors declare that no funds, grants, or other support were received during the preparation of this manuscript.

## Authors’ Contributions

- **Parand Shariat Rad:** Conceptualization, Methodology, Writing - Original Draft.
- **Mohammad Mehdi Hatami & Mohammad Amin Amani:** Formal analysis, Data Curation, Validation.
- **Shayan Salehi & Mohammad Hossein Sharif & Mohammad Javad Gharegozlou:** Investigation, Writing - Review & Editing.
- **Mohammad Gharib Salehi:** Supervision, Project administration, Final approval.

## Acknowledgment

The authors would like to acknowledge that the MeDeep artificial intelligence platform evaluated in this study was built upon the underlying architecture of CheXagent, a vision-language foundation model developed for chest X-ray interpretation. We extend our appreciation to the researchers who developed the CheXagent model and its associated large-scale training dataset, CheXinstruct.

## Notes

### Competing Interest Statement

The authors have declared no competing interest.

### Author Declarations

Ethics Committee of Kermanshah University of Medical Sciences gave ethical approval for this work (Approval No: IR.KUMS.MED.REC.1405.053).

